# The Clinimetric Profile of 11 Generic Quality Indicators for Appropriate Antibiotic Use in Hospitalized Patients in Indonesia

**DOI:** 10.1101/2023.11.22.23298934

**Authors:** Rahajeng N. Tunjungputri, Khie Chen Lie, Adityo Susilo, Freddy C. Nainggolan, Chyntia O.M. Jasirwan, Rudy Hidayat, Dyah Purnamasari, Robert Sinto, Erni J. Nelwan

## Abstract

**Background:** One of the efforts in antibiotic stewardship for limiting antibiotic resistance is the assessment of the quality of antibiotic use. To date, the quality and appropriateness of antibiotic use in Indonesia, a low-middle-income non-European country, have not yet been assessed using proven quality indicators (QIs). One recommended tool to measure appropriate empiric antibiotic use in hospitalized patients is the generic QIs reported by van den Bosch *et. al*., which demonstrated good validity and generalizability.

**Objective:** The aim of this study is to assess the clinimetric profile of 11 generic QIs based on van den Bosch *et. al*. in admitted adult patients receiving empiric antibiotics in Indonesian hospitals.

**Methods:** This is an observational study with a cross-sectional design conducted at two government referral hospitals in Jakarta, Indonesia, from August 1, 2022, to February 2, 2023. Adult inpatients who received empiric antibiotics due to suspected infection for ≥24 hours were included in the study.

**Results:** In 500 inpatients from 2 hospitals, all QIs demonstrated good measurability with less than 10% of missing data. Ten QIs showed good applicability of >10%. Four QIs show good performance, while six QIs have significant potential for improvement (two blood cultures are obtained before empiric antibiotic treatment; culture sample is obtained from location suspected of infection; antibiotic is switched from intravenous to oral when clinically possible; antibiotics are stopped when there is no evidence of infection; a guideline is available and renewed every three years; and a guideline is adjusted to the local resistance pattern) making them priority targets for interventions to enhance the quality of antibiotic use in Indonesia. All QIs display good inter-observer reliability, and no moderate or strong correlations are found between QIs.

**Conclusion:** The clinimetric assessment of QIs is imperative before their implementation in any setting different from their country of origin. Of the 11 generic QIs, 10 demonstrated good reliability and applicability at two hospitals in Indonesia. Future intervention studies can utilize these generic QIs to measure improvement in the appropriateness of antibiotic use.

## Introduction

The increasing antimicrobial resistance (AMR), accompanied by a decline in the discovery of new antibiotics, has become a global crisis.^1^ The emergence of multidrug-resistant organisms, including including extended spectrum beta-lactamases (ESBL) producing Enterobacterales, extensively drug-resistant (XDR) carbapenemase-producing *Klebsiella pneumoniae* and other Enterobacterales, as well as *Pseudomonas aeruginosa* and pan-drug-resistant (PDR) *Acinetobacter baumannii* has major impact on the mortality and morbidity of infections.^2^ Antibiotic stewardship is defined as coordinated interventions designed to improve and measure the appropriate use of antibiotics by promoting the selection of the optimal drug regimen including dosing, duration of therapy, and route of administration.^3,4^ Robust antibiotic stewardship has several benefits for clinicians, such as reducing antibiotic prescriptions, lowering the cost of managing infection cases, and reducing the number of infections caused by multidrug-resistant organisms (MDRO).^3,5,6^

One aspect of antibiotic stewardship is improving the quality of antibiotic use. A method to systematically assess the quality of antibiotics use is by using Quality Indicators (QIs) developed from various international guidelines and literature. QIs are often the result of a rigorous, multidisciplinary process.^7–9^ However, they cannot automatically be implemented in daily practice for all settings, such as in low- or middle-income countries. The validation of QIs in everyday clinical practice is crucial before using them to measure the quality and appropriateness of antibiotic use.^10^

Indonesia is a low-middle-income country in Southeast Asia with a population of 275 million and substantial inequalities across the country.^11^ In 2019, AMR accounts for the fourth largest cause of deaths just under cardiovascular diseases, malignancy and diabetes.^12^ The National Action Plan for AMR exists, with antimicrobial stewardship programs in place and national health scheme aiming to achieve universal health care.^13,14^ However, challenges remain with limited data on AMR epidemiology and inappropriate antibiotic use. Appropriate prescribing was estimated at 33.5% using the Gyssens method.^14,15^ The quality of antibiotic prescribing practices in the country are not yet assessed with the implementation of QIs.^15^ With many hospitals lacking the infrastructure to adequately measure process, outcome and structural indicators, such as insufficient training and infectious diseases and microbiology expertise, there is a need for QIs that are easily applicable and can soundly measure the quality of antibiotic use.

Published sets of QIs often were particular to a specific clinical syndrome^16–19^ or patient population in the hospital.^8,20^ A recommended set of 11 generic QIs is one reported by van den Bosch et al.,^21^ which was reported to have high validity and generalizability for use in various hospital settings and infectious diseases. This study was aimed to assess the clinimetric profile (measurability, applicability, performance, improvement potential, inter-QI correlation, and Kappa coefficient) of the QIs based on van den Bosch *et. al.* for evaluating the appropriateness of antibiotic use in adult inpatients in 2 referral hospitals in Indonesia.

## Methods

### Setting, study population and data collection

This was a multicenter observational study in which we measured the clinimetric profiles of 11 generic QIs by van den Bosch et. al.^21^ in 2 government referral hospitals, with Hospital 1 being an academic, national tertiary referral hospital and Hospital 2 being a nonacademic, secondary referral hospital. The medical records were electronic and paper-based in Hospital 1 and Hospital 2, respectively. Both hospitals had antibiotics stewardship programs at the time the study was conducted, and were on the national health scheme, which fully covered expenses for treatment as well as microbiology culture and sensitivity testing. The patients were included between July 2022-November 2022.

To include a representative group of patients, we identified in each hospital on all departments, admitted adult patients who received empiric antibiotics for more than 24 hours because of suspected infection on the day of patient screening, using the point prevalence survey (PPS). For all admitted patients, those on antibiotics were screened and recruited in the study when meeting the study criteria. The hinclusion criteria were patients age ≥18 years old, had been started on systemic antibiotics at least 24 hours earlier, and having suspicion of bacterial infection. The exclusion criteria were patients on prophylactic antibiotics, and those who started antibiotics from a different hospital. For all included patients, clinical data and laboratory tests of the entire treatment course with antibiotics, from the start of antibiotic therapy until hospital discharge, were retrospectively extracted from medical and nursing records. To rule out prophylaxis, antibiotics had to be started at least 24 hours before inclusion. All data needed to calculate IQ were obtained by trained general practitioners. The data were extracted uniformly and entered into an electronic case report form (eCRF). This study is approved by the ethics committee of the Faculty of Medicine, Universitas Indonesia.

### Quality indicators and definitions

We used the set of 11 generic QIs for appropriate antibiotic use developed by van den Bosch *et al.,* using a systematic modified Delphi method.^21,22^ The first 9 indicators are process indicators, while the last 2 indicators are structural indicators.^21^ All 11 QIs were tested for their clinimetric profiles in the two hospitals in our study. The list of the 11 generic QIs and their definitions are listed in **Table 1**. The criteria for safe switching within 72 h of treatment were as follows: (i) the patient was hemodynamically stable; (ii) no fever; (iii) normal white blood cell count; (iv) able to take oral medication; and (v) functioning gastrointestinal tract, without signs of malabsorption. A safe switch was not possible in cases of meningitis, intracranial abscesses, endocarditis, mediastinitis, *Legionella pneumonia*, exacerbations of cystic fibrosis, inadequately drained abscesses with empyema, severe soft tissue infections such as group A streptococcal infections, infections of foreign bodies, *Staphylococcus aureus* or *Pseudomonas aeruginosa* bacteremia, liver abscesses with empyema, osteomyelitis or arthritis, or chemotherapy-related neutropenia.^23^

**Table 1.** The list of the 11 generic QIs and their definitions.

| No | QI | Applicability |  | Performance |  |
| --- | --- | --- | --- | --- | --- |
|  |  | Numerator | Denominator | Numerator | Denominator |
| 1 | **Empirical systemic antibiotic therapy should be prescribed according to the national guideline.** | Total number of patients who started with empirical systemic antibiotic therapy and an infection clinical syndrome presents in the antibiotics guideline. | Number of patients receiving empiric antibiotics. | Number of patients who started with empirical systemic antibiotic therapy according to the antibiotics guideline. | Total number of patients who started with empirical systemic antibiotic therapy and an infection clinical syndrome presents in the antibiotics guideline. |
| 2 | **Before starting systemic antibiotic therapy, at least two sets of blood cultures should be taken.** | Total number of patients who started with systemic antibiotic therapy. | Number of patients receiving empiric antibiotics. | Number of patients in whom at least two samples of blood cultures were taken before systemic antibiotic therapy was started. | Total number of patients who started with systemic antibiotic therapy. |
| 3 | **When starting systemic antibiotic therapy, specimens for culture from suspected sites of infection should be taken as soon as possible, preferably before antibiotics are started.** | Total number of patients who started with systemic antibiotic therapy. | Number of patients receiving empiric antibiotics. | Number of patients in whom cultures from suspected sites of infection were taken within 24 hr after the systemic antibiotics were started. | Total number of patients who started with systemic antibiotic therapy. |
| 4 | **An antibiotic plan should be documented in the case notes at the start of systemic antibiotic therapy.** | Total number of patients who started with systemic antibiotic therapy. | Number of patients receiving empiric antibiotics. | Number of patients for whom an antibiotic plan was documented in the case notes. | Total number of patients who started with systemic antibiotic therapy. |
| 5 | **Systemic antibiotic therapy should be switched from i.v. to oral antibiotic therapy within 48-72 hr on the basis of the clinical condition and when oral treatment is adequate.** | Total number of patients with i.v. antibiotics for 48-72 hr, in whom changing to oral antibiotic therapy on the basis of the clinical condition was indicated. | Number of patients receiving empiric antibiotics. | Number of patients with i.v. antibiotics for 48-72 hr, in whom changing to oral antibiotic therapy on the basis of clinical conditions was performed. | Total number of patients with i.v. antibiotics for 48-72 hr, in whom changing to oral antibiotic therapy on the basis of the clinical condition was indicated. |
| 6 | **Empirical antibiotic therapy should be changed to pathogen-directed therapy if culture results become available.** | Total number of patients with empirical systemic antibiotics, whose culture became positive. | Number of patients receiving empiric antibiotics. | Number of patients with empirical therapy whose culture became positive and changing to pathogen-directed therapy was performed correctly. | Total number of patients with empirical systemic antibiotics, whose culture became positive. |
| 7 | **Dose and dosing interval of systemic antibiotic therapy should be adapted to renal function.** | Total number of patients who started with systemic antibiotic therapy which should be dosed according to renal function and who had an unknown or compromised renal function | Number of patients receiving empiric antibiotics. | Number of patients with a compromised renal function with a dosing regimen adjusted to renal function. | Total number of patients who started with systemic antibiotic therapy which should be dosed according to renal function and who had an unknown or compromised renal function |
| 8 | **Therapeutic drug monitoring should be performed when the therapy duration is >3 days for aminoglycosides and >5 days for vancomycin.** | Total number of patients who received aminoglycosides for >3 days and/or vancomycin for >5 days. | Number of patients receiving empiric antibiotics. | Number of these patients with at least one serum drug level measurement. | Total number of patients who received aminoglycosides for >3 days and/or vancomycin for >5 days. |
| 9 | **Empirical antibiotic therapy for presumed bacterial infection should be discontinued based on the lack of clinical and/or microbiological evidence of infection. The maximum duration of empirical systemic antibiotic treatment should be 7 days.** | Total number of patients who started empirical systemic antibiotic therapy, but lacked clinical and/or microbiological evidence of infection. | Number of patients receiving empiric antibiotics. | Number of patients whose empirical antibiotic therapy was discontinued within 7 days. | Total number of patients who started empirical systemic antibiotic therapy, but lacked clinical and/or microbiological evidence of infection. |

### Analysis

Collected data were entered into a database using the SPSS 25.0 for Windows® (SPSS Inc., Chicago, IL, USA). A descriptive analysis was performed for each indicator. Applicability represents the percentage of evaluated patients who had the necessary information to calculate the QIs. Performance was defined as the percentage of patients who adhered to the QIs. The potential for improvement for each IQ was calculated as 100% minus the performance score and was used to identify areas for improvement in the antibiotic prescriptions.

Fifty samples were evaluated simultaneously by 2 investigators, to calculate inter-observer reliability for the performance scores of all QIs. The rate of agreement was calculated and expressed as kappa coefficients. Correlation between performance scores of the quality indicators were calculated using Spearman coefficient.

## Results

The study included 500 subjects from Hospital 1 (n=317) and Hospital 2 (n=173). The screening of patients resulting in the included subjects are presented in **Figure 1**. The patient characteristics are presented in **Table 2**. The most common clinical syndrome was lower respiratory tract infection (n=281, 56.2%), with sepsis found in 14.4% of subjects. Other infections included brain and meningeal infections, acute gastroenteritis, exit site infections, osteoarticular infections, surgical wound infections, central line-associated bloodstream infections, mediastinitis, infected implants, upper respiratory tract infections, and lung abscesses. The distribution of wards in the two hospitals are presented in **Figure 2A**. Two or more infection diagnoses were present in 28,6% of subjects. All antibiotics (n=500, 100%) were initially administered intravenously.

**Figure 1.**
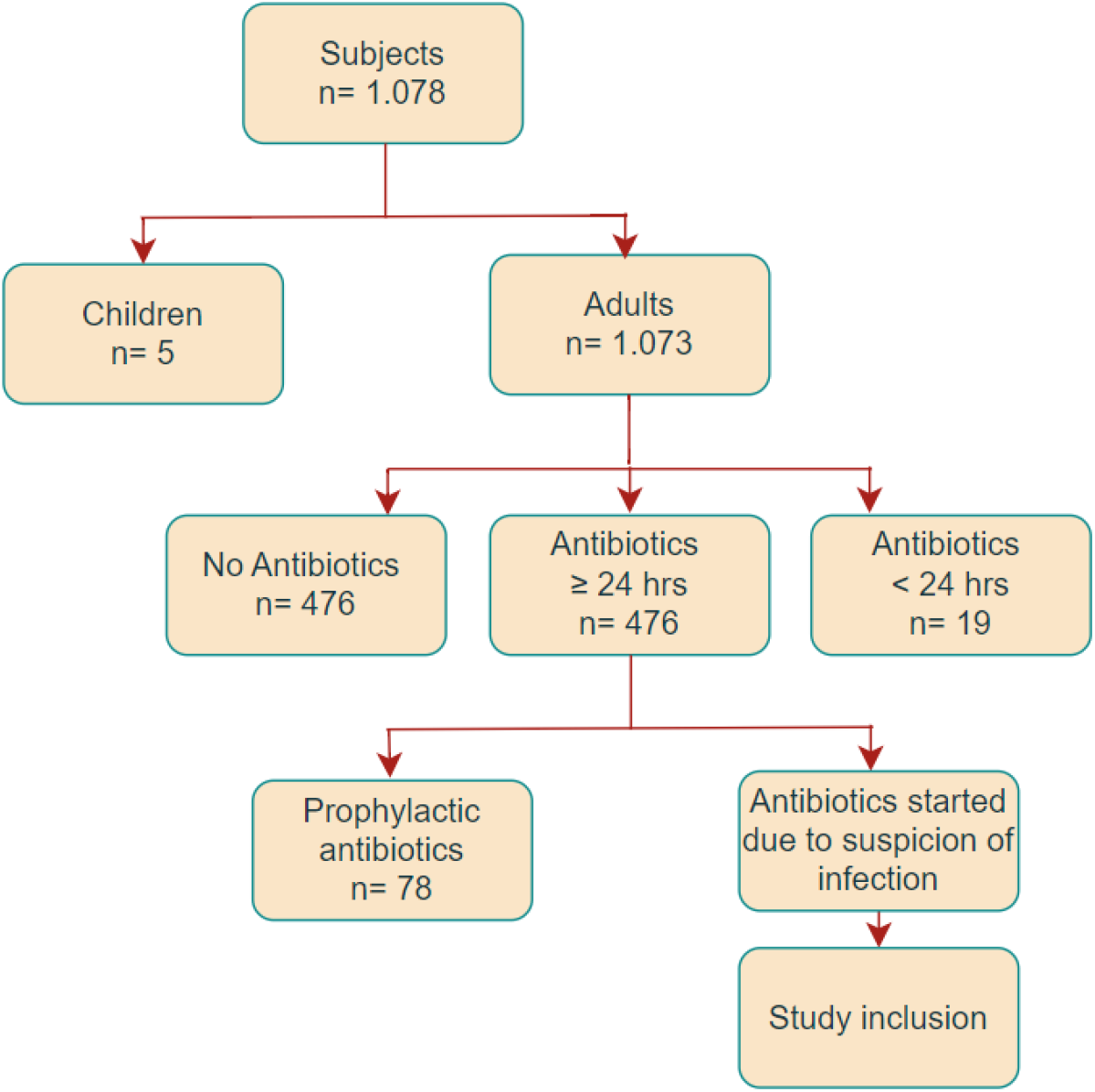
Subject recruitment. Patients identified during the screening process.

**Figure 2.**
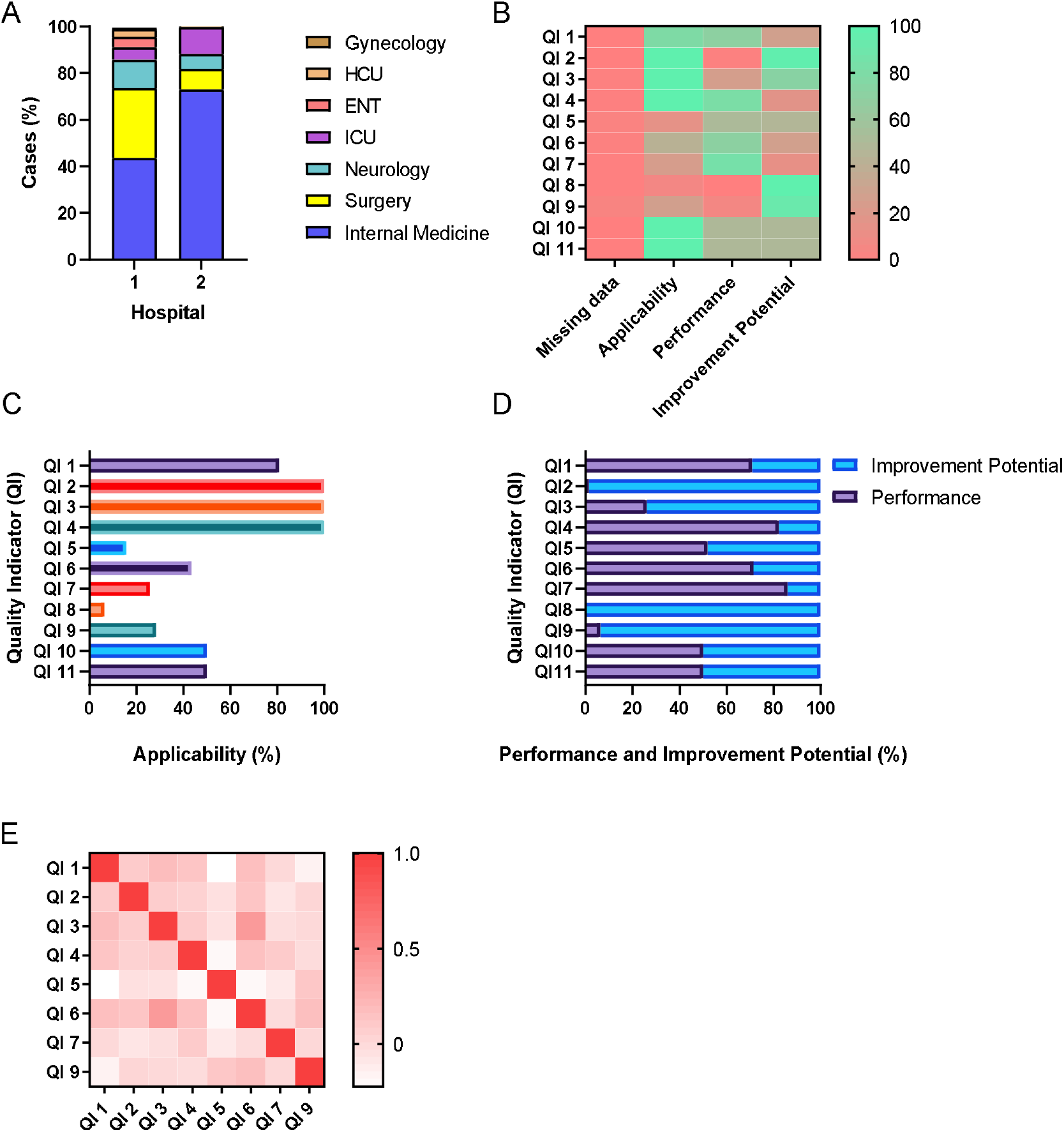
The clinimetric profile of 11 generic quality indicators of appropriate antibiotic use for hospitalized patients. (A) Distribution of wards in 2 hospitals where subjects were included into the study. (B) Clinimetric profile heat map showing percentage clinimetric properties for all QIs. (C) The applicability of QIs in percentage. (D) The performance and improvement potentials of QIs in percentage. Heat map showing the Spearman correlation coefficient between all applicable QIs at the patient level.

**Table 2.** Clinical characteristics.

| **Characteristics** | **Number** |
| --- | --- |
| **Sex** |  |
| Male (n, %) | 238 (48) |
| Female (n, %) | 262 (52) |
| **Age (median, IQR)** | 53 (42-63)* |
| **Clinical syndrome** |  |
| Lower respiratory tract infection (n, %) | 281 (56,2) |
| Urinary tract infection (n, %) | 110 (22) |
| Skin and soft tissue infection (n, %) | 73 (14,6) |
| Sepsis (n, %) | 72 (14,4) |
| Biliary tract infection (n, %) | 15 (3) |
| Intraabdominal infection (n, %) | 10 (2) |
| Two or more infection diagnosis (n, %) | 143 (28,6) |
| Others (n, %) | 31 (6,2) |

\*Data is shown in median (interquartile range, IQR) for data with nonnormal distribution. HCU, *high care unit*. ICU, *intensive care unit*.

The clinimetric profile data are shown as heat map in **Figure 2B**, with data presented in **Table 3**. All QIs had good measureability in both hospitals, with missing data <10%. Ten QIs demonstrated good applicability (**Figure 2C**), except for QI 8 (therapeutic drug monitoring for aminoglycosides after >3 days and vancomycin after >5 days) with the applicability score of only 6,4%. Similar to the measurability scores, the applicability scores between the two hospitals in this study was also comparable.

**Table 3.**
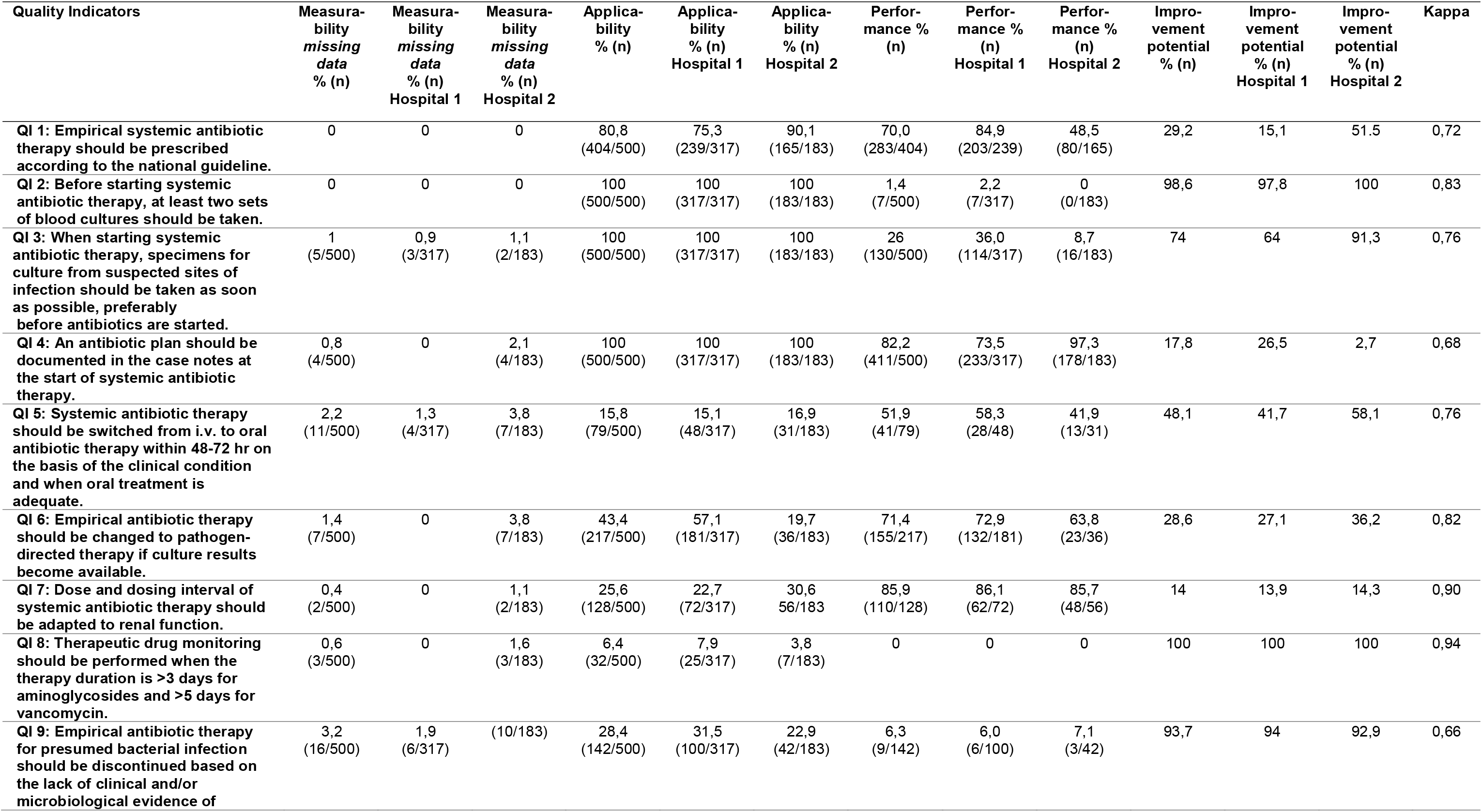
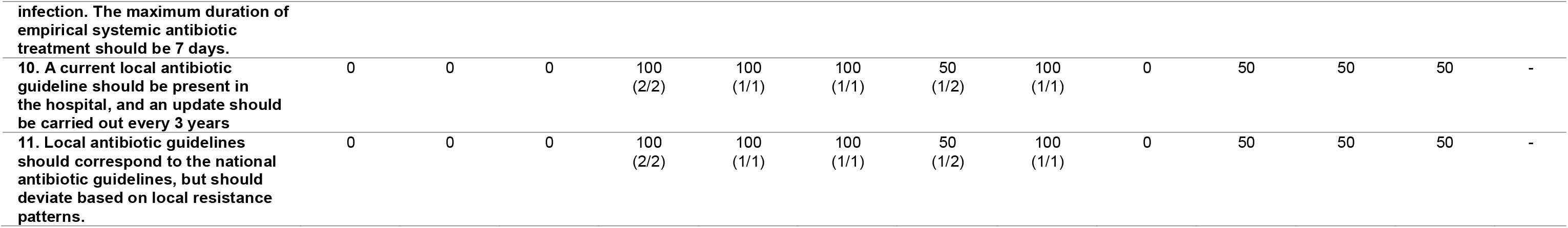

Both hospitals exhibited variability in QI performance scores (**Figure 2D**). Hospital 1 had markedly higher performance scores than Hospital 2 for QI 1 (guideline concordance; 84.9% vs. 48.5%) and QI 3 (cultures obtained from suspected infection sites; 36.0% vs. 8.7%). In contrast, Hospital 2 had a higher performance score for QI 4 (documentation of antibiotic therapy plans at initiation; 97.3% vs. 73.5%). Among all applicable QIs, QI 2 on obtaining 2 sets of blood culture had the lowest performance score. Only 1,7% patients had two sets of blood culture taken, with 17% had one set of blood culture taken. Only 22 out of 72 subjects (30,5%) with sepsis had their blood culture taken.

In this study, four patient-level QIs were found to have improvement potential scores above 30%: QI 2 (two blood cultures taken before antibiotic initiation, 98.6%), QI 3 (cultures obtained from suspected infection sites, 74%), QI 5 (switching from IV to oral antibiotics if clinically feasible, 48.1%), and QI 9 (discontinuation of empirical therapy if no clinical and/or microbiological evidence of infection, 93.7%). Additionally, QI 10 and QI 11 regarding hospital-level guidelines had a 50% potential for improvement. In this study, all QIs had a Kappa coefficient >0.6, and there was weak correlation between QIs, making it impossible to reduce the number of QIs for implementation.

## Discussion

The need for appropriate antibiotic use to curb the growing antimicrobial resistance can be met with applying a set of QIs as part of an effective antimicrobial stewardship program. We herewith describe 10 generic QIs for appropriate antibiotic use in hospitalized patients with sound clinimetric properties applicable for daily use in Indonesia, with 6 QIs demonstrated improvement potential for prioritization of intervention. To the best of our knowledge, this is the first study identifying generic QIs with sound clinimetric profile for appropriate antibiotic use in all types of infections in Indonesia. Previously, performance properties of QIs for antimicrobial use have been assessed, but mostly only in European countries.^7,9,17,18,21,24^ Farida *et. al.*^16^ previously developed QIs for antibiotic use and measure their clinimetric properties in Indonesia, but only for pneumonia.

Interestingly, 10 out of the 11 QIs in this study demonstrated sound applicability, in contrast to only 7 reported by van den Bosch *et. al.*^21^ in their validation study. Therapeutic drug monitoring (TDM) QI showed low applicability score due to the low use of vancomycin and aminoglycoside in our setting. This finding is also in line with that from van den Bosch *et. al.*^21^ and Arcenillas *et. al.*^25^ The QI was subsequently dismissed from our performance assessment. Indeed, TDM in Indonesia has not yet become part of routine clinical practice. This was largely due to the cost and expertise required,^26^ and the relative lack of need therein, as shown by this study. We would subsequently recommend this QI to also be dropped from being used elsewhere as 3 studies including ours^21,25^ have demonstrated its low applicability both in high- and middle-income countries.

The improvement potential measures the sensitivity of a QI to detect variation in quality of care between hospitals and within a hospital. High-performance scores with low variation between hospitals render a QI insensitive and less useful in clinical practice because it has low potential for improvement. An improvement potential of >30% is considered sufficient for focusing intervention strategies on improving antibiotic use quality in a hospital. Four QIs at the patient level as well as two QIs at the hospital level were shown to have good improvement potential in this study. The prioritized QIs were: QI 2 (two blood cultures taken before antibiotic initiation), QI 3 (cultures obtained from suspected infection sites), QI 5 (switching from IV to oral antibiotics if clinically feasible), and QI 9 (discontinuation of empirical therapy if no clinical and/or microbiological evidence of infection) which focus on diagnostic and de-escalation practices.

Process indicators are useful in determining the impact of antimicrobial stewardship. Beyond capturing the state of antibiotic use, the implementation of QIs may also aid in measuring how far an intervention impacts the quality antibiotic use.^27,28^ Measurement in the quality of antibiotic use has long been part of the routine practice in Indonesia. However, to date, the widely used audit method in Indonesia is the Gyssens flow chart, which was developed by Gyssens *et. al*.^29^ in 1992, both for clinical and research purposes.^15,30^ Compared to the Gyssens method, these 11 generic QIs by van den Bosch *et. al.* provided more insights into antibiotics prescribing practices, which includes assessments of: 1) processes prior to antibiotic prescribing (collection of 2 sets of blood culture samples and samples from suspected sites of infection), 2) documentation in the antiobiotic prescribing plan, 3) safe switch from iv to oral route; deescalation process which include 4) dose adjustment according to renal function (which captures the safety aspect of antibiotic use) and 5) stopping empiric antibiotic therapy when appropriate, as well as the availability of updated local antibiotics guidelines that are adapted to local antimicrobial resistace pattern at the hospital level.

QI 2 (obtaining 2 sets of blood culture) performed extremely low in this study at 1,4%; whereas when 1 blood culture was obtained the adherence number increased to 17%. This finding was in line with that from Sinto *et. al.*^31^ which found that at least 13% of patients had their blood sample taken once for culture. The low number of blood culture utilization in our setting may be due to the lack of clear guidelines applied for identifying the indications of blood culture sampling, despite their availability.^32–34^ Furthermore, the QI with the biggest discrepancy in improvement potential among the two hospitals in this study is QI 1 (adherence to guideline). This may be affected by the fact that Hospital 2 with the lower performance score did not have their own antibiotics guideline, and without the national antibiotic guideline being used formally at the hospital. Therefore, decisions to start or choose which antibiotics were left for the clinicans, based on national/international guidelines, or to their own discretion.

Indeed, barriers to antimicrobial stewardship in Indonesia still included the factors of antibiotic decision-making by clinicians. A study using questionnaires by Limato *et. al*.^35^ reported these factors include insufficient application of antimicrobial stewardship principles and microbiology services, the lack of confidence in prescribing decisions, and defensive prescribing due to diagnostics uncertainties, fear of patient deterioration, or because the patient insisted. A wide variation exists between hospitals, departments, work experience and medical hierarchy.^11,15^ The utilization of QIs could potentially overcome these issues by introducing them as actionable points. The 11 generic QIs in this study would provide clear guidance on appropriate antibiotic prescribing and empower clinical decision-making. Kallen *et. al.*^8^ developed a set of four indicators for appropriate antibiotic use in the ICU which were introduced as actionable QIs, together with an implementation toolbox which includes checklists and improvement strategies. Such strategies would render QIs not only useful for quality assessment, but also as a method of intervention.

This study has several strengths. The selected 11 generic QIs were obtained through a multidisciplinary Delphi-RAND methodology,^21,22^ and were previously validated in a large group of patients,^21,25^ albeit exclusively in a European setting. Secondly, this study provides an insight into the clinimetric profiling of these QIs in a diverse, lower-middle income country, demonstrating its robustness. Thirdly, the study showed that assessment of these 11 QIs can be performed by a small number of people trained to collect this information from either an electronic or paper-based medical records, with good interobserver reliability, which supports its wide and ease of implementation.

The potential limitations of this study are the involvement of two centers which are both located in Jakarta, the capital city of Indonesia. This would mean that the two hospitals have more similarities than its eastern or outer islands counterparts in Indonesia. Secondly, we had intended to include subjects in the ICU, as previous studies^21,25^ did not measure these QIs in the aforementioned population. However, we did not have a large number of patients originating from the ICU, and thus the specific applicability of these QIs in the ICU patients remains to be determined.

In conclusion, the clinimetric assessment of QIs is imperative before their implementation in any setting different from their country of origin. Of the 11 QIs from van den Bosch *et. al.,* 10 demonstrated good reliability and applicability at the two hospitals in Indonesia, a low-middle-income non-European country. Interventions to improve the quality of antibiotic use can be focused on the six QIs with the greatest potential for improvement. Future intervention studies can utilize these generic QIs to measure improvement in the appropriateness of antibiotic use.

## Data Availability

All data produced in the present study are available upon reasonable request to the authors.

## Notes

### Competing Interest Statement

The authors have declared no competing interest.

### Funding Statement

This study was funded by PUTI Q1 grant from Universitas Indonesia.

### Author Declarations

The ethics committee of the Faculty of Medicine, Universitas Indonesia gave ethical approval for this work.

### Summary of Updates

There is correction in the institutions of all authors. There is no correction in the content of the manuscript.

